# Pan-resistome characterization of uropathogenic *Escherichia coli* and *Klebsiella pneumoniae* strains circulating in Uganda and Kenya isolated from 2017-2018

**DOI:** 10.1101/2020.03.11.20034389

**Authors:** Arun Gonzales Decano, Kerry Pettigrew, Wilber Sabiiti, Derek Sloan, Stella Neema, Joel Bazira, John Kiiru, Hellen Atieno Onyango, Benon Asiimwe, Matthew T. G. Holden

## Abstract

**Background:** Urinary tract infection (UTI) develops after a pathogen adhered to the inner lining of the urinary tract. Cases of UTIs are predominantly caused by several Gram-negative bacteria and account for high morbidity in the clinical and community settings. Of greater concern are the strains carrying antimicrobial resistance (AMR)-conferring genes. The gravity of UTI is also determined by a spectrum of other virulence factors. This study represents a pilot project to investigate the burden of AMR among uropathogens in East Africa.

**Methods:** We examined bacterial samples isolated in 2017-2018 from in- and out-patients in Kenya (KY) and Uganda (UG) that presented with clinical symptoms of UTI. We reconstructed the evolutionary history of the strains, investigated their population structure and performed comparitive analysis their pangenome contents.

**Results:** We found 55 *Escherichia coli* and 19 *Klebsiella pneumoniae* strains confirmed uropathogenic following screening for the prevalence of UTI virulence genes including *fimH, iutA, feoA*/*B*/*C, mrkD* and *foc*. We identified 18 different sequence types in *E. coli* population while all *K. pneumoniae* strains belong to ST11. The most prevalent *E. coli* sequence types were ST131 (26%), ST335/1193 (10%) and ST10 (6%). Diverse plasmid types were observed in both collections such as Incompatibility (IncF/IncH/IncQ1/IncX4) and Col groups. Pangenome analysis of each set revealed a total of 2,862 and 3,464 genes comprised the core genome of *E. coli* and *K. pneumoniae* population, respectively. Among these are AMR determinants including fluoroquinolone resistance-conferring genes *aac*(3*)-Ib-cr* and other significant genes: *aad, tet, sul1, sul2*, and *cat*, which are associated with aminoglycoside, tetracycline, sulfonamide and chloramphenicol resistance. Accessory genomes of both species collection were detected several β-lactamase genes, *bla*_CTX-M_, *bla*_TEM_ and *bla*_OXA_ *or bla*_*NDM*._Overall, 93% are multi-drug resistant in the *E. coli* collection while 100% of the *K. pneumoniae* strains contained genes that are associated with resistance to 3 or more antibiotic classes.

**Conclusions:** Our findings illustrate the abundant resistome and virulome repertoire in uropathogenic *E. coli* and *K. pneumoniae*, which are mainly disseminated via clonal and horizontal transfer, circulating in the East African region. We further demonstrate here that routine genomic surveillance is necessary for high-resolution bacterial epidemiology of these important AMR pathogens.

## Introduction

Antimicrobial resistance (AMR) has raised alarms as a global health threat. AMR is often fuelled by misuse and abuse of antibiotics including self-medication (Michael et al., 2014; Rather et al. 2017) and unrestricted access to antimicrobial drugs (Nathan and Cars 2014, French 2010, Read and Woods 2014), and is further accelerated by industrialization, poor waste disposal and hygiene levels. AMR pathogens are frequently detected in food, clinical and environmental settings in East Africa and despite facing broad challenges, significant efforts have recently been put in place to curb AMR in East African countries. For instance, Kenya (KY) has adapted the National Action Plan that incorporates One Health measures to prevent AMR and is highly supported by multiple governmental policies (NAPCAR 2017). Similarly, an extensive evaluation of the AMR situation in Uganda (UG) was assessed by the Uganda National Academy of Sciences (UNAS) supported by the Global Antibiotic Resistance Partnership (GARP)-Uganda (UNAS 2015). High prevalence of multi-drug resistant bacteria particularly extended-spectrum beta-lactamase (ESBL)-producing strains is significantly recorded in both countries.

Urinary tract infection (UTI) develops after a pathogen’s adherence to the inner lining of the urinary tract. UTIs occur among patients of all age groups and account for high morbidity in the clinical and community settings (Flores-Mireles 2015). Following binding within the urinary tract, uropathogens either cause asymptomatic or commensal connection, or severe disease. About 1% of the population have asymptomatic bacteriuria (ABU), wherein a pathogen (≥10_5_ cfu mL_−1_) inhabits the tract without eliciting mucosal host response (Kunin et al. 1962; Lindberg et al. 1975). Infections in the lower urinary tract region e.g. cystitis, are recognized by symptoms such as dysuria. Successful virulent strains can induce pyelonephritis where rapid immune response is mobilised via cytokine secretion and influx of immune cells. UTIs are either uncomplicated or complicated. Uncomplicated UTI cases are usually observed in patients who are otherwise healthy, while complicated UTIs are diagnosed in compromised patients e.g. if have anatomical or functional anomalies in their urinary tract or are under long-term catheterization (Najar et al. 2009). Treatment of these complicated UTI cases is often confounded by AMR uropathogens usually caused by Gram-negative bacteria (Levison and Kaye 2013). Uncomplicated UTIs are frequently caused by uropathogenic *Escherichia coli* (UPEC) while complicated cases might be caused by several pathogens such as *Proteus mirabilis, Providencia stuartii, Morganella morganii, Klebsiella pneumoniae and Pseudomonas aeruginosa* (Flores-Mireles et al. 2015). Recurrent UTI cases are also common, particularly when urinary tract anomalies linger, or treatment failed to kill resistant bacteria (Nicolle LE 2000), leading to more severe type of infections. Due to the lack of active investigation of UTI cases in East Africa, particularly in the comunity, access to accurate data can be challenging.

An increasing number of studies have employed whole genome sequencing (WGS) and analyses for disease surveillance in both hospital and community settings (Croucher et al. 2015; Goswami et al. 2018). The high resolution genotyping that WGS provides can be used to investigate and describe the population structure, and evolutionary history of the isolates, as well as tracing their spread. Outbreaks have been robustly detected and described using high-throughput methodologies designed for bacterial pathogens (Halachev et al. 2014; Lee et al. 2018; Ludden et al. 2020). Comprehensive AMR gene databases and prediction tools are also available that help assess AMR gene content in whole genomes with high accuracy (Feldgarden et al. 2019). Here, we used WGS to investigate the prevalence of AMR-conferring genes in *E. coli* and *K. pneumoniae* isolated from urine samples taken from patients in rural areas of KY and UG that presented UTI-like symptoms. We further explored their phylogenetic relationships with other currently circulating African and global strains. This study represents a pilot project of the HATUA consortium. HATUA stands for Holistic Approach to Unravel Antibiotic Resistance in East Africa and the team is comprised of researchers from different disciplines that aim to tackles the main drivers of AMR among uropathogens in East Africa.

## Methods

### Study design and Patient recruitment

A total of N=150 bacterial isolates were obtained from patients in KY (n=91) and UG (n=59) presenting UTI-like symptoms, as part of a larger study. Ethical Review Board of University of St Andrews ethical approval, Approval code MD14548 and KY (KEMRI/SERU/P00112/3865) approved verbal consent taken from all the patients. Important patient data such as name, age, gender, location was recorded, and unique identification number were assigned to each patient.

### Library preparation and whole genome sequencing

Bacterial genomic DNA for the isolates were extracted using the QIAxtractor (Qiagen, Valencia, CA, USA) according to the manufacturer’s instructions. Library preparation was conducted according to the Illumina protocol and sequenced (96-plex) on an Illumina MiSeq X platform (Illumina, San Diego, CA, USA) using 250 bp paired-end reads.

### Read library quality control, mapping and de novo genome assembly

Illumina MiSeq read libraries were rid of sequencing adapters and ambiguous bases using Fastp (Chen at al. 2018). Sets that passed the quality filtering were *de novo* assembled using Unicycler v4.6 (Wick et al. 2017) pipeline in normal mode to merge contigs.

The read libraries were mapped to reference sequences using SMALT v7.6 (http://www.sanger.ac.uk/resources/software/SMALT/) and the resulting SAM files were converted to BAM format, sorted and PCR duplicates removed using SAMtools v1.19 (Li et al. 2009). Strain TOP52_1721_U1 (Wiley et al. 2019) was used the reference genome for the *K. pneumoniae* samples while the strain EC958 (Forde et al. 2014) was employed as the reference sequence for the *E. coli* population.

### Species identification, sero- and sequence typing, genome annotation and screening for UTI virulence/AMR genes

Prediction of bacterial species was done by uploading the assemblies on PathogenWatch website (https://pathogen.watch), which runs Speciator (https://gitlab.com/cgps/mash-speciator) for its species assignment. Speciator employs Mash (Ondov et al. 2016) to identify the most identical strain (≥90% identity) in a reference collection of complete genomes found in the NCBI RefSeq database (https://www.ncbi.nlm.nih.gov/refseq/). The strains are then grouped according to their species designation and were screened for UTI pathogen determinants. Multi-locus sequence typing was performed by running SRST2 v.0.2.0 (Inouye et al. 2014) based on the Achtman scheme (Achtman et al. 2012) for *E. coli* and Pasteur (Diancourt et al. 2010) for *K. pneumoniae* isolates. Antigenic (O polysaccharide and H flagellin) profiles of *E. coli* samples were identified by employing Serotypefinder v.2.0 (https://cge.cbs.dtu.dk/services/SerotypeFinder/) at 85% ID threshold and 60% minimum length.

Genome composition of the draft assemblies was assessed using Prokka v.1.10 (Seemann 2014). AMR genes were identified by aligning the genome sequences to the 2,158 gene homolog subset of the Comprehensive Antibiotic Resistance Database (CARD) v. 3.0.8 (https://card.mcmaster.ca/). Plasmid and replicon typing was done by comparing the genomes against the PlasmidFinder database v. (Carattoli et al. 2014) at 99% identity threshold.

### Bacterial sample collection and Antimicrobial Susceptibility Testing

To determine concordance between the AMR gene content and sample phenotype, antibiotic testing and phenotypic detections of ESBL were performed by disc diffusion methods on a subset of n=16 isolates from KY; the tests are done according to CLSI, 2016 guidelines. Isolates were examined for the insusceptibility to 9 different classes of antibiotics including Penicillin (ampicillin (AMP)), Penicillin + β-lactamase inhibitors (ampicillin-clavulanic acid (AMC)), Chloramphenicol (Chloramphenicol (CHL)), Sulfonamide (Trimethoprim-sulfamethoxazole (SXT)) and Quinolones (nalidixic acid (NA)) and Fluoroquinolone (Ciprofloxacin (CIP)). Resistance to ESBL Cephalosphorins was also assessed by testing the strains with Ceftriaxone (CRO), Ceftazidime (CAZ), cefotaxime (CTX) and Cefepime (FEP) (Supplementary Table 1).

**Table 1.** UTI virulence marker genes present in the pangenome of N=55 *E. coli* and N=19 *K. pneumoniae* isolates. Proportion of the samples containing the gene are shown in count of strains with gene over the total strains and % values.

| Gene | Protein Product | Strain count and % in *E. coli* collection | Strain count and % in *K. pneumoniae* collection |
| --- | --- | --- | --- |
| *fimH* | Type 1 fimbrial D-mannose specific adhesin | 56/56 (100) | 20/20 (100) |
| *feoA/B/C* | Fe(2+) transport protein A/B/C | 56/56 (100) | 20/20 (100) |
| *entB* | Enterobactin synthase component B | 56/56 (100) | 20/20 (100) |
| *focA* | Formate transporter | 56/56 (100) | 20/20 (100) |
| *zapA* | Cell division protein | 56/56 (100) | 20/20 (100) |
| *satP* | Succinate-acetate/proton symporter | 56/56 (100) | 20/20 (100) |
| *chuR* | Anaerobic sulfatase-maturing enzyme | 56/56 (100) | 19/20 (95) |
| *mrkD* | Type 3 fimbrial adhesin | 0 | 20/20 (100) |
| *hlyE* | Hemolysin E | 45/56 (80.4) | 2/20 (10) |
| *fyuA* | Pesticin receptor | 41/56 (73.2) | 3/20 (15) |
| *iutA* | Ferric aerobactin receptor | 25/56 (44.6) | 2/20 (10) |
| *papD* | Import of P pilus subunits into the periplasm | 26/56 (46.4) | 2/20 (10) |
| *papA* | Fimbrial major pilin protein | 23/56 (41.1) | 0 |
| *kpsT* | Polysialic acid transport ATP-binding protein | 15/56 (26.8) | 0 |
| *pic* | Serine protease pic autotransporter | 3/56 (5.4) | 0 |

### Phylogenetic reconstruction

Phylogenetic relationships and sequence variations between the samples were determined by constructing phylogenetic trees based on their chromosomal SNPs; MGEs were further excluded using an internal script. Non-recombinant SNPs were determined using ClonalframeML v. 1.12 (Didelot and Wilson 2015) and were used to create a maximum-likelihood midpoint-rooted phylogeny using RAxML v8.0.19 (Stamatakis 2014) using a General Time Reversible + gamma (GTR+G) substitution model with 100 bootstraps. Phylogenies were visualized using iToL (https://itol.embl.de/) and FigTree v1.4.3 (http://tree.bio.ed.ac.uk/software/figtree/).

### Pangenome analyses

The resulting annotation files from Prokka v.1.10 (Seemann 2014) were used as the basis for generating a pangenome for each species set. This step was done by running Roary v3.11.2 (Page et al. 2015) with a 100% BLAST v2.6.0 identity threshold using the MAFFT v7.3 setting (Katoh and Standley 2013). Pangenome outputs were also used to assess the accessory genome composition of each bacterial population and as basis for constructing core genome phylogenies.

## Results

### Patient and bacterial strain profiles

A collection of N=100 strains, n=81 *E. coli* and n=19 *K. pneumoniae* were isolated from urine of different patients admitted or visiting rural hospitals in Kenya and from clinics in the countryside of Uganda.

### *Genomic and pangenomic characterization confirmed the virulence factors present in uropathogenic* E. coli *and* K. pneumoniae

A subset (n=55) from the total n=81 *E. coli* and all n=19 *K. pneumoniae* were confirmed uropathogenic following a thorough characterization of their pangenome contents. One thousand one hundred forty-four (1,144) and 3,464 core genes were found across the strains in *E. coli* and *K. pneumoniae* populations, respectively. These include known UTI virulence markers that are responsible for urinary tract (mucosal) surface binding (type 1 fimbrial adhesin-coding *fimH*) and colonization (*mrkD*; *K. pneumoniae* only), iron (Fe(2+)) transport (*feoA/B/C*), enterobactin synthase production (*entB*), formate transport (*focA*), cell division (*zapA*), succinate-acetate/proton symport (*satP*), anaerobic sulfatase-maturation (*chuR*; found in 100% and 95% of *E. coli* and *K. pneumoniae*, respectively). Other important virulence genes were also found, albeit not conserved among all the isolates: *iutA* (ferric aerobactin receptor: 44.6% in *E. coli*, 10% in *K. pneumoniae*), *papA* (fimbrial major pilin protein: *E. coli* only (41%)), *papD* (import of P pilus subunits into the periplasm: 44.6% in *E. coli*, 10% in *K. pneumoniae*), *hlyE* (hemolysin E: 80.4% in *E. coli*, 10% in *K. pneumoniae*), *fyuA* (pesticin receptor: 73.2% in *E. coli*, 15% in *K. pneumoniae*), *kpsT* (polysialic acid transport ATP-binding protein: *E. coli* only (26.8%)) and *pic* (serine protease pic autotransporter: *E. coli* only (5.4%); Table 1).

### Prevalence of AMR genes in E. coli and K. pneumoniae uropathogens from KY and UG

All n=55 *E. coli* and n=19 *K. pneumoniae* isolates harboured type 1 fimbrin. Among the UPEC, *fimH30* was the most common allele, followed by *fimH41*; n=4/55 samples had type *fimH22* and n=2/55 singleton were found with *fimH22*.

We further detected multiple AMR-conferring elements in the genomes of the two species collections. Alignment of the sequences against CARD v.3.0.7 with 98-100% identity revealed that the n=55 *E. coli* (n=31 from KY, n=24 from UG) were detected with the ciprofloxacin-conferring gene, *marA*. Majority (n=47/55) were also aminoglycoside resistant and harbours either *aadA* or *aac(6’)-Ib/(3’)-Ib* alleles or both. Only n=11/55 were not detected with resistant genes for ESBL cephalosphorins. Of the n=44/55 that produce ESBLs, n=10/44 had *bla*_CTX-M_ (allele type 15 or 88), n=24/44 had *bla*_TEM_(type 30/2/220) and n=9/44 had *bla*_OXA-1/140_ and n=2/44 (both from UG) had all 3 ESBL genes. Sulfonamide resistance was widely observed, n=39 had either *sul1* only, *sul2* only or *sul3* only, or both *sul1* and *sul2*. Tetracycline resistance gene, *tet(A)* was present in n=34 of 55. Twenty-four (N=24/55) contained macrolide resistance-conferring *mphA*, n=9/55 (n=6 from KY, n=3 from UG) harboured *catB3* and were chloramphenicol resistant. Ninety-five percent (95%, n=52/55) had at least one gene that codes for efflux pump proteins with n=4/55 having *yojI-pmrF-emrR-bacA*-*acrS/B/E-msbA-evgA-kdpE-mdtP-eptA* cassette and n=1/55 containing a mixture of *yojI, pmrF, emrR, bacA, acrS/B/E, msbA, evgA, kdpE, mdtP, eptA, emtK, cpxA* (Table 2a). Two KY isolates (71 and 72) were found to have the fluoroquinolone resistance-conferring gene *aac(3’)-Ib-cr* while its variant *aac(6’)-Ib-cr* was present in UG isolates BN19, BN38 and BN44.

All n=19 *K. pneumoniae* isolates were resistant to aminoglycosides and had either *aac(6’)-30/aac(6’)-Ib’/7/10-aadA9* (n=10/19) or *aac(6’)-Ib7-aadA9* (n=1/19) combination. N=18/19 are potentially sulfonamide insusceptible and contained either *sul*1 only (n=2/19), *sul*2 only (n=15/19) or both (n=1). The β-lactamase *bla*_LEN_ gene is present in all but the BN7 strain, with *bla*_LEN-4/6_ (n=15 from KY) or *bla*_LEN-3/4/5/6_ alleles. BN7 was also the only susceptible isolate against ESBL cephalosphorins. The rest are ESBL producers: n=15/19 had *bla*_SHV-28_, *bla*_CTX-M-15_, *bla*_OXA-1/140_ and *bla*_NDM-1_, n=1/19 were observed with the *bla*_SHV-28-CTX-M-15-OXA-1/140_ combination and n=1/19 had *bla*_SHV-28_, *bla*_CTX-M-15/88_ only. All n=3 strains from UG had *tet(B)* and *dfrA* (17 or 27 allele type); these strains also contained efflux pump-expressing genes: *yojI, pmrF, emrR, bacA, acrB, msbA, evgA, kdpE, mdtP, eptA, emtK and cpxA*. N=2/19 (BN14 and BN16 from UG) had ciprofloxacin-resistance gene, *marA*. Only the strain 90 from KY was not resistant to phenicols, while the rest were detected with the *cat* gene, specifically, *catB3* (Table 2b). Overall, 80% of our E. coli uropathogens had ESBL genes (n=15 strains from UG and n=29 from KY) and 93% of these UPEC are MDR, while all the *K. pneumoniae* isolates are MDR and only n=1 out of the total n=19 (95%) are ESBL.

### Population structure of KY and UG Uropathogens

The UPEC collection was polyclonal. Eighteen (18) different sequence types were identified in the UPEC population (Achtman scheme). The most prevalent MLST sequence types were ST131 (n = 17/55, 31%), ST335 and ST1193 (n=6/55, 11%) and ST10 (n= 4/55, 7%). These sequence types were usually associated with UTI cases (Nicolas-Chanoine et a. 2014; Afset et al. 2008; Yamaji et al. 2018); the globally disseminated ESBL-ST131 stood out to be the most dominant ST. Other clones were also observed: n=3 ST73, n=2 each from ST155, ST410, ST6161 and ST162, and singletons from ST44, ST48, ST165, ST167, ST212, ST448, ST617, ST648 and ST2163; n=2 strains from UG (BN2 and BN48) were unclassified (Figure 1a, Table 2a). *E. coli* isolates from UG belong to 15 STs and were thus more diverse compared to those collected from KY, which belong to only 6 STs (Figure 1a, Table 2a). This difference in diversity is consistent with the number of serotypes found in UG relative to those from KY: Ugandan strains belong to 20 different O:H antigen combinations while the KYn ones were found to be have 9 O:H types.

**Figure 1.**
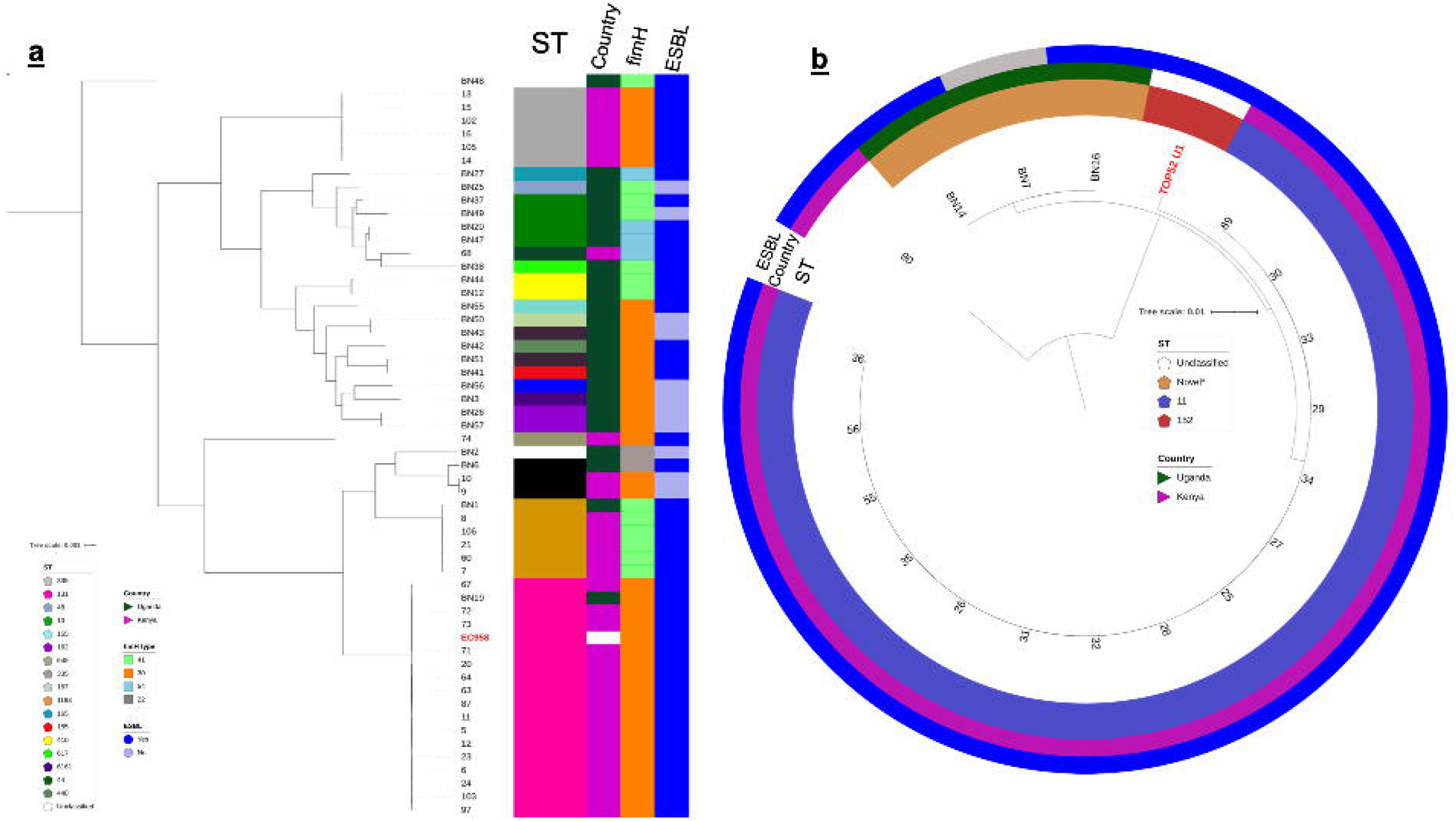
Maximum likelihood phylogenies of core genomes of *E. coli* (a) and *K. pneumoniae* (b) uropathogens isolated from KY and UG; reference genomes are in red and font. The mid-point rooted phylograms was constructed using 2,862 and 3,464 core genes from *E. coli* and *K. pneumoniae* populations, respectively and visualized with iTol. The coloured strips adjacent to the *E. coli* phylogeny represent (from left to right) the sequence type (ST), country of origin, type of *fimH* allele and the ESBL status of each strain. The coloured rings around the *K. pneumoniae* phylogenetic tree indicate the ST, country of origin and the ESBL status of each isolate. The scale bar indicates the number of SNP distance from the reference.

Fifteen out of nineteen (n=15/19) *K. pneumoniae* isolates from KY belong to ST11 (Pasteur scheme); n=3/19 UG had no defined sequence types (BN14: 0b8e, BN16; 67b2, BN7: 6b6f) and formed their own clade (Table 2b; Figure 1b).

### Plasmid characterization

Genome assemblies of the KY and UG uropathogens were screened for the presence and type of plasmids using PlasmidFinder v. N=47/55 in the *E. coli* collection were found with at least one plasmid. IncFIA was consistently found in n=10 had both *Inc*FIA and *Inc*FII, n=9 contained *Inc*FIA, *Inc*FII, *Col*156 types, n=1 was detected with *Inc*FIA, *Inc*FII and *Inc*Y only or *Inc*I only and *Inc*FII-*Inc*FIA-*Inc*X4 plasmid combinations.

All the samples from the *K. pneumoniae* collection were found with at least one plasmid type. *Inc*FII-*Inc*FIB-*Inc*R is the most common combination and is found among n=15 isolates, while n=2/19 was found with *Inc*R, *Inc*FII, *Inc*FIA, *Col* and *Inc*X4. Notably, the strain 90 from KY had the *bla*_CTX-M-1_ gene-carrying plasmid *Inc*N and BN7 from UG had the *bla*_NDM_-associated *Inc*R.

## Discussion

We assessed the prevalence of AMR characteristics among uropathogenic *E. coli* and *K. pneumoniae* circulating in East African region using WGS. We recruited out-patients that presented UTI-like symptoms from rural areas in KY and UG, which represents a limitation of our sample collection. The lack of point-of-care diagnostic tool such as the use of dipstick test also contributed to some difficulties in our screening. This is evidenced by a high level of contaminants that comprised of strains that do not contain UTI determinants. Nevertheless, our *in silico* predictions using whole genome analysis revealed alarming rates of ESBL-producing and MDR strains in both our UPEC and *K. pneumoniae* collections, which reiterates the great necessity for effective interventions to curb their spread.

Our results firmly indicate a high diversity among *E. coli* uropathogens, which was more evident in samples taken from UG rather than KY. Strains that belong to the same clonal group had <200 core SNPs from each other. This rich genetic diversity is consistent with those observed in other isolates collected from rural or semi-rural communities of low/middle-income countries (Salinas et al. 2019; Kimata et al. 2005; Montealegre et al. 2020). The widely-disseminated UTI-causing clones ST131, ST335 and ST10 were common among our *E. coli* strains and dominated our Kenyan collection. This is unsurprising as these ST’s are reported to be circulating globally (Ben Zakour 2016; Sanjar et al. 2015; Day et al. 2019). What’s remarkable is the detection of emerging clones such as ST1193 and ST617 that were unusually associated with UTI (Valenza et al. 2019; Mhaya et al. 2019) albeit observed in hospital settings. UPEC strains from UG are even more alarming as they represent higher number of unusual or novel UTI clones i.e. ST155, ST448 and ST162 with potentially higher virulence levels (Castellanos et al. 2017; Choudhury 2018; Zhang et al. 2019) compared to those globally-known STs.

*Bla*_CTX-M_ genes were present in 40% of our UPEC collection and in all but one *K. pneumoniae* strain (95%). Notably, the *bla*_CTX-M-15_ gene that confers resistance to last-resort antibiotics was found in high levels in both countries. This gene was detected with other ESBL determinants, *bla*_TEM_ and *bla*_OXA-1_ in *E. coli* and with *bla*_NDM_ in *K. pneumoniae*, concordant with those in uropathogens found from the Middle East (Alqasim et al. 2018) and Asia (Zhang et al. 2015), among others. Consistent with previous findings in other African regions, *tet* genes in this study were also detected alongside ESBL genes *bla*_CTX-M-15_, *bla*_OXA-1_ and *bla*_TEM_ in n=30/55 *E. coli* and with *bla*_LEN-3/4/5/6_ among n=3/19 *K. pneumoniae* (Mbelle et al. 2019; Adefisoye et al. 2016) which stipulates their co-selection and co-transmission in KY and UG. The presence of these genes in the identified plasmid-asscociated contigs suggest that the mode of transfer may have been plasmid-mediated.

Taken together, we underline in this pilot study the high frequency of AMR determinants associated with resistance to common antibiotic classes among *E. coli* and *Klebsiella pneumoniae* in East Africa, with specific focus on MDR and ESBL-producing strains from KY and UG. We further demonstrate that routine genomic surveillance is necessary for high-resolution investigation of bacterial epidemiology especially in less represented regions. Our findings have significant implications on improving interventions that aim to address the strong presence of AMR pathogens that cause UTI particularly in low/middle-income countries.

## Data Availability

The genome data is in the process of being submitted to the public sequences databases

## Supplementary Materials

Supplementary Table 1. Metadata of *E. coli* and *K. pneumoniae* strains isolated from urine samples including Antibiotic Sensitivity Test results of n=16 isolates.

## Acknowlegments

We thank Prof. Andy Lynch from the Mathematical Institute at the University of St. Andrews for his helpful feedback on the manuscript.

## Figures and Tables

Table 2. Genomic characteristics of uropathogenic *E. coli* (a) and *K. pneumoniae* strains (b) isolated in this study for AMR-associated genes and plasmid replicon types. NF means not found; Yes means the strain is either ESBL-producing or MDR and No means the sample is either non-ESBL or non-MDR. (View or download from Figshare, DOI: https://figshare.com/articles/Table_2a_and_2b/11965455).

